# Epidermal Growth Factor in the Brain: A Promising Biomarker for Depression

**DOI:** 10.1101/2024.08.01.24311271

**Authors:** Shu-xian Xu, Honggang Lyu, Mian-mian Chen, Kun Li, Lihua Yao, Chao Wang, Xin-hui Xie, Zhongchun Liu

## Abstract

**Background:** This study aimed to examine the neurotrophic factors secreted from brain in depression by analyzing astrocyte-derived extracellular vesicles (ADEVs) isolated from plasma, and to explore the causal relationship between the expression of neurotrophic factors in the brain and depression.

**Methods:** A total of 40 patients with treatment-resistant depression (TRD) and 35 matched healthy controls (HCs) were recruited at baseline, and 34 TRD patients completed the post-electroconvulsive therapy (ECT) visits. The concentrations of five neurotrophic factors in ADEVs were measured. A correlation analysis was performed between neurotrophic factors in ADEVs and neurogenesis marker doublecortin (DCX) in neuron-derived extracellular vesicles (NDEVs). Subsequently, Mendelian randomization (MR) study and cell experiments were conducted.

**Results:** Our findings revealed a decrease in the level of epidermal growth factor (EGF) in ADEVs among TRD patients, with an increase observed post-ECT. The corrected area under the curve for EGF were larger than those for other neurotrophic factors: 0.99 (95% CI: 0.98-1.00). MR suggested that decreased expression levels of the *EGF* gene in the cortex constitute a risk factor for depression. We observed a positive correlation between the levels of EGF in ADEVs and DCX in NDEVs. Subsequently, cell experiments suggested that EGF can activate EGF receptor (EGFR) to trigger the PI3K-Akt pathway, participating in the promotion of DCX.

**Conclusions:** This study provides the *in vivo* evidences supporting that a reduction in EGF levels in the central nervous system could potentially contribute to depression and serve as a biomarker for it. Additionally, the EGF/EGFR signaling pathway may be involved in regulating early neurogenesis traits in depression.

## Introduction

Major depressive disorder (MDD) is among the most prevalent mental disorders, marked by significant and enduring depression, loss of interest, and anhedonia^1,2^. It is also a leading cause of disability worldwide and contributes substantially to the global burden of disease^3^.

Various hypotheses have been proposed to explain the underlying causes of depression^4,5^. Among them, the neurotrophic hypothesis of depression suggests that MDD-related synaptic and brain changes are primarily due to disrupted neurotrophic support^5^. Neurotrophic factors are of major importance because it modulates the plasticity and neurogenesis, inhibits cell death cascades and increases cell survival proteins that are responsible for proliferation and maintenance of central nervous system (CNS) neurons^4,6^.

Astrocytes, known for their critical role in regulating neurotrophic factors and gliotransmitters that maintain brain homeostasis^7,8^, may potentially contribute to the neuropathology of depression^9,10^. Astrocytes produce various neurotrophic factors, including brain-derived neurotrophic factor (BDNF), glial cell line-derived neurotrophic factor (GDNF), epidermal growth factor (EGF), fibroblast growth factor (FGF), insulin-like growth factor (IGF-1), vascular endothelial growth factor (VEGF), and nerve growth factor (NGF), all of which are implicated in the etiology and treatment of depression^11,12^. Among them, EGF is a protein that could be valuable for the early diagnosis and treatment assessment of depression. For instance, MDD patients with the rs11569017-TT genotype showed low plasma levels of EGF when compared to controls^13^. MDD patients exhibit reduced EGF receptor *ErbB3* expression, which increases and correlates with clinical improvement following 12 weeks of antidepressant treatment ^14^. This suggests that investigating variations in the *EGF* gene and their receptor expression could serve as a clinical predictor of recovery in depression.

Unfortunately, research on other neurotrophic factors in depression remains limited, with inconsistent findings. Peripheral blood samples, used in many previous studies^15^, might not accurately reflect brain neurotrophic factor levels due to their widespread expression in various tissues^16–18^. Investigating these factors directly within the brain is likely crucial for a deeper understanding of depression. The difficulty of directly measuring neurotrophic factor levels in the CNS hinders the acquisition of *in vivo* molecular evidence in patients with MDD. However, recent breakthroughs in isolating brain-derived extracellular vesicles (EVs) offer a revolutionary non-invasive approach for assessing CNS status *in vivo*. These EVs hold immense promise as a “liquid biopsy” to evaluate molecular alterations within the brain^19–21^.

In this study, we initially found a decrease in the level of EGF in astrocyte-derived extracellular vesicles (ADEVs) among treatment-resistant depression (TRD) patients compared to controls, with an increase observed post-electroconvulsive therapy (ECT). Subsequently, a Mendelian randomization (MR) study was conducted to explore the causal effect of *EGF/EGF receptor (EGFR)* gene expression on depression from a genetic perspective. As we also observed a positive correlation between EGF levels in ADEVs and the early neurogenesis marker doublecortin (DCX) in neuron-derived extracellular vesicles (NDEVs). Cell experiments using SH-SY5Y cells were then performed to investigate the relationship between EGF and DCX.

## Methods

### Participants details

This observational longitudinal cohort was conducted at Renmin Hospital of Wuhan University in accordance with the Declaration of Helsinki^22^ as our previous study^23,24^. The Human Ethics Committee approved the study protocol. Patients or their legal guardians provided informed consents. This report follows the STrengthening the Reporting of Observational studies in Epidemiology (STROBE) statement^25^.

A total of 40 patients with TRD and 35 age-, and sex-matched healthy controls (HCs) were recruited. Subjects were excluded if they had comorbidity with other major psychiatric disorders. The HCs were required to be in good physical health, with no personal or first-degree family history of psychiatric disorders.

Thirty-four patients with TRD completed the post-ECT visits. Six milliliters of blood were drawn at baseline (T1), and 24 hours after the final ECT session (T2) to mitigate acute treatment effects. Cohort details and ECT procedures can be found in supplementary materials and our previous studies^23,24^.

### Isolation and confirmation of ADEVs

We employed different isolation methods for ADEVs used in protein measurements and cell treatments. A two-step ADEVs protocol based on our prior work^24^ was used for protein analysis. In contrast, ADEVs for cell treatment underwent ultracentrifugation (UC)^26^ to isolate the total plasma EVs, followed by the same immunoprecipitation method to isolate ADEVs. This approach avoided potential contamination and cellular effects associated with polymer precipitation methods commonly used in clinical settings. Confirmation of ADEV isolation was performed using transmission electron microscopy (TEM), nanoparticle tracking analysis (NTA), and western blotting. Please refer to the supplementary materials and our previous studies^24^ for more details.

### Protein measurements in ADEVs

The levels of CD81, an EV marker, was measured using enzyme-linked immunosorbent assay (ELISA) kit (American Research Products Inc., Catalog# CSB-EL004960HU). Neurotrophic factors BDNF, FGF-2, GDNF, EGF and NGF-β in ADEVs were measured using 5-Plex Human ProcartaPlex™ Panel (Thermo Fisher Scientific).

### Causal genetic evidence from Mendelian randomization

A MR study was conducted to validate the association between the neurotrophic factors identified above and depression from a genetic perspective. By utilizing genetic variants as instrumental variables, MR can provide more robust causal inference between exposures and outcomes compared to traditional observational studies, thereby mitigating common confounding factors such as residual confounding, reverse causation bias, and measurement error. In this study, we leveraged brain-tissue specific expression quantitative trait loci (eQTL) data and genome-wide association study (GWAS) summary statistics on depression to perform two-sample MR (**sFigure 1**). Wald ratio and inverse variance weighting (IVW) were employed as the primary analytical methods in the main analysis. Additionally, sensitivity analyses were conducted. Details are provided in the supplementary materials.

**Figure 1.**
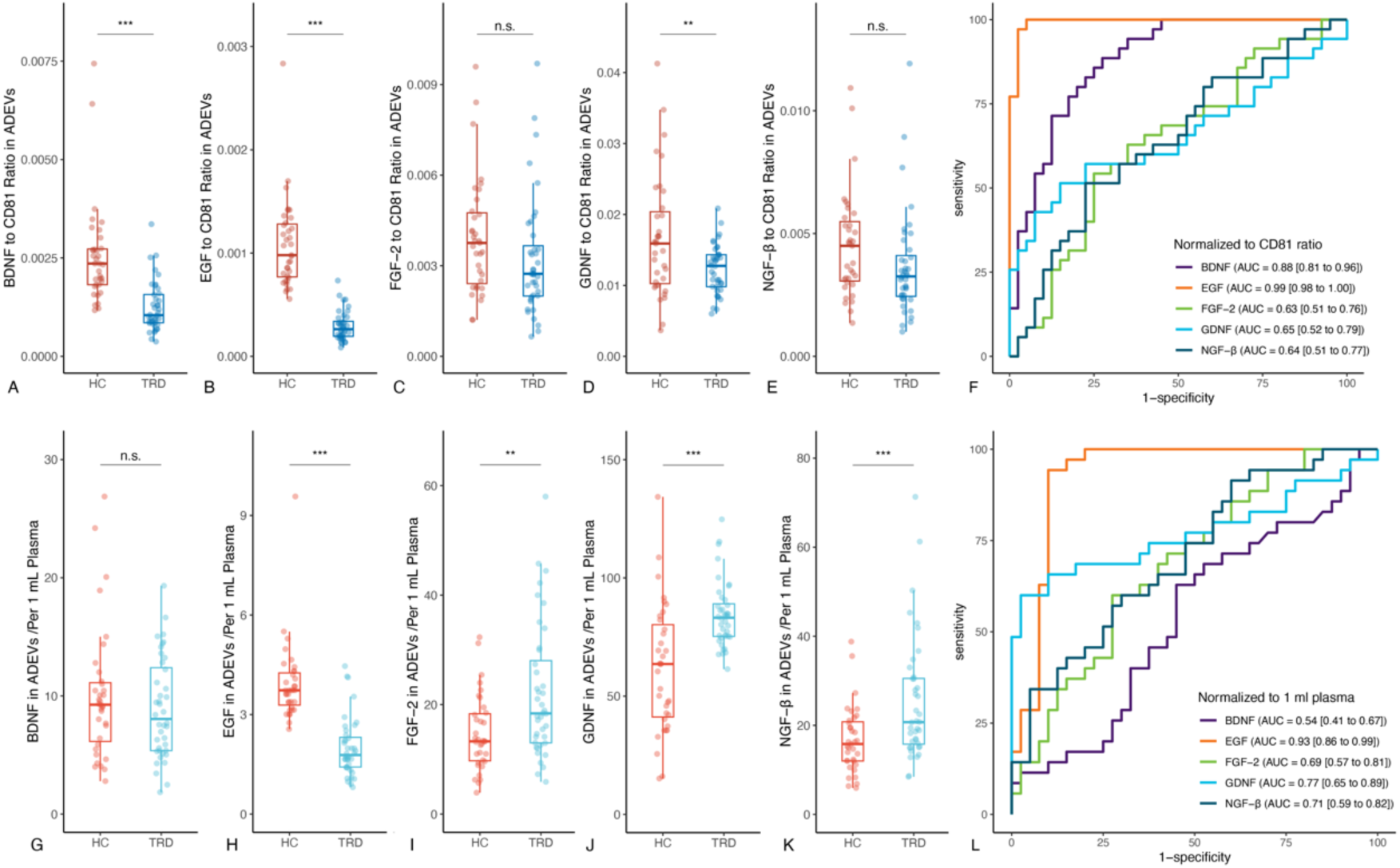
Comparisons of target protein concentrations in astrocyte-derived extracellular vesicles (ADEVs) between treatment-resistant depression (TRD) and healthy control (HC) group at baseline. Note: n.s.: not significant; \**p*_false discovery rate (FDR)_ < 0.05; \*\**p*_FDR_ < 0.01; \*\*\**p*_FDR_ < 0.001.

### Culture and treatments of SH-SY5Y cells

The SH-SY5Y cell line was obtained from Wuhan Procell Biotech Co., Ltd., China. These cells were cultured in basic DMEM/F-12 medium (Gibco, Catalog# C11330500BT) supplemented with 10% fetal bovine serum (FBS, WISENT, Catalog# 086-150) and 100 U/ml Penicillin-Streptomycin (Gibco, Catalog# 15140122) under 5% CO_2_ at 37 °C. The medium was changed every 2-3 days, and when the cell density reached 80-90%, they were passaged at a 1:3 ratio.

Next, the well-growing SH-SY5Y cells were seeded in a 6-well plate at a density of 6 *10^5^ cells per well with 2.5 ml medium for 24 hours. All cells used in the experiment were in the logarithmic growth phase. The cells were divided into two groups: the HC group (treated with ADEVs from 6 HCs) and the TRD group (treated with ADEVs from 6 patients with TRD). The EGF levels in ADEVs from HCs were higher than those in TRD. ADEVs were diluted with culture medium to a final concentration of 20 μg/ml. After incubating with the respective agents for 24 hours, the cells were harvested for subsequent analysis.

Furthermore, we performed another experiment, the well-growing SH-SY5Y cells were seeded in a 6-well plate at a density of 6 *10^5^ cells per well with 2.5 ml medium for 24 hours. All cells used in the experiment were in the logarithmic growth phase. Recombinant human EGF protein was diluted to a final concentration of 10 ng/ml to treat SH-SY5Y cells. The control group received an equal volume of growth medium instead of the EGF solution. After 24 hours of treatment, the cells were harvested for analysis.

### Western blotting

Previous studies found that EGF binds to its receptor EGFR, activating the PI3K-Akt and MAPK-Erk pathways which are crucial for neurogenesis in depression^27–29^. Therefore, we analyzed the proteins in these two pathways.

The following proteins were detected in SH-SY5Y cells: DCX (Abcam, Catalog# ab207175), GAPDH (Huabio, Catalog# HA721131), EGFR (Affinity, Catalog# AF6043), Phospho-EGFR (Tyr1068) (Affinity, Catalog# AF3045), Akt (Cell Signaling, Catalog# 9272S), Phospho-Akt (Ser473) (Cell Signaling, Catalog# 4060S), PI3K p85 (Affinity, Catalog# AF6241), Phospho-PI3K p85 (Affinity, Catalog# AF3241), Erk1/2 (Cell Signaling, Catalog# 9102S), Phospho-Erk1/2 (Thr202/Tyr204) (Cell Signaling, Catalog# 4370T).

### Statistical analyses

For the clinical data, in the main analysis of protein levels in ADEVs at baseline, following the MIESV 2018^30^, the outcomes were defined as the ratio of protein concentrations to CD 81 concentrations. General linear models (GLMs) were used to examine differences between two groups. In the GLMs, we set the outcomes as dependent variables and group as the independent variable, while controlling for covariates for sex and age. To represent the differences between the two groups, we used model-fitted estimated marginal means (EMMs) and their corresponding 95% confidence intervals (CIs). Additionally, to ensure robustness, a sensitivity analysis was conducted, with protein concentration levels per 1 ml plasma set as dependent variables. Receiver operating characteristic curves (ROCs) and corrected area under the curve (AUC) were employed to further explore the potential of these neurotrophic factors in ADEVs as biomarkers. In the exploratory analysis, Spearman coefficient was used to examine the correlation between EGF in ADEVs and DCX in NDEVs (as reported in our previous report^23^). For the comparisons between pre- and post-ECTs (paired longitudinal data), paired *t*-tests were used, with Cohen’s *d* and its associated 95% CIs utilized to quantify effect sizes. Similar to the comparisons at baseline, a sensitivity analysis was conducted with the concentrations of protein levels per 1 ml plasma as dependent variables. False discovery rate (FDR) correction (Benjamini and Hochberg method) was employed to mitigate false positive outcomes. In comparisons of data from cell experiments, the Wilcoxon tests were employed. A two-tailed *p*-values less than 0.05 was considered significant. All statistical analyses were performed using R version 4.1.0 (R Project for Statistical Computing) within Rstudio version 1.4.1106 (Rstudio).

## Results

### EGF levels in ADEVs represent a promising potential biomarker for depression

Among 40 patients with TRD, six patients either did not undergo ECT treatment or did not complete the post-ECT visit. Detailed baseline characteristics of the participants was provided in **sTable 1**.

At baseline, the level of EGF in the TRD group was significantly lower than in the HC group. This effect was consistent across different normalization methods. Changes in other neurotrophic factors were more variable and yielded different outcomes depending on the normalization method used (**Table 1** and **Figure 1**). The AUCs for EGF were larger than those for other neurotrophic factors, with values of 0.99 (95% CI: 0.98-1.00) and 0.93 (95% CI: 0.86-0.99), respectively (**Figure 1**). This indicates that EGF in ADEVs has a strong discrimination ability. Additionally, the EGF level in ADEVs showed a positive correlation (*ρ* = 0.38, *p* < 0.001) with the DCX level in NDEVs (**sFigure 2**).

**Figure 2.**
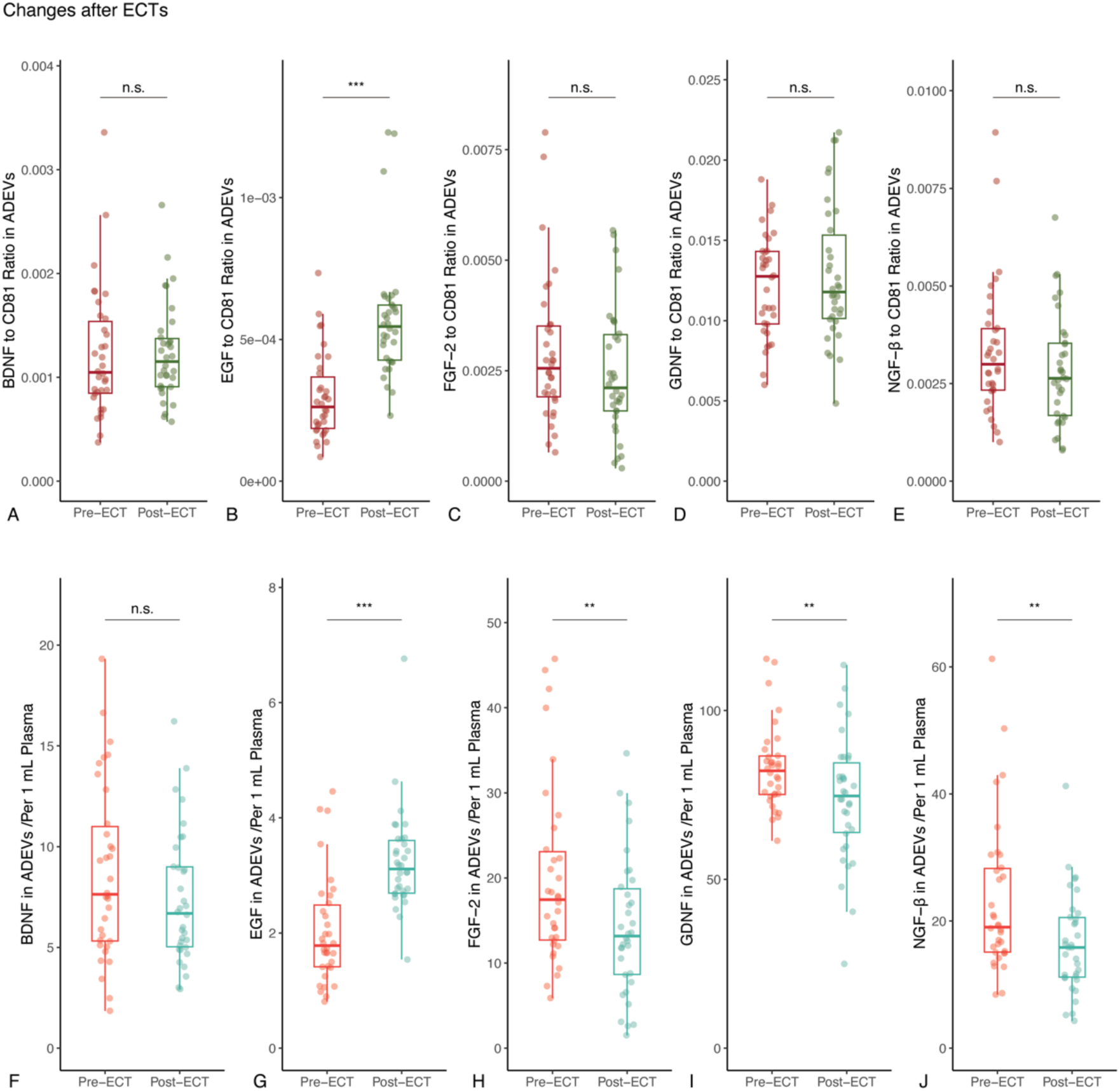
Changes of target protein concentrations in ADEVs between pre- and post-ECT in patients with TRD. Note: n.s.: not significant; \**p*_false discovery rate (FDR)_ < 0.05; \*\**p*_FDR_ < 0.01; \*\*\**p*_FDR_ < 0.001.

**Table 1.** Baseline comparisons of neurotrophic factors in ADEVs between TRD and HC groups.

| variable | HC group (EMM (95% CI)) | TRD group (EMM (95% CI)) | Difference (95% CI) | *p* | *p<sub>FDR</sub>* |
| --- | --- | --- | --- | --- | --- |
| Main analysis: Normalized to CD81 ratio |  |  |  |  |  |
| BDNF | 0.0025 (0.0022 to 0.0029) | 0.0012 (0.0009 to 0.0015) | -0.0013 (-0.0018 to -0.0009) | <0.001 | <0.001 |
| EGF | 0.0011 (0.001 to 0.0012) | 0.0003 (0.0002 to 0.0004) | -0.0008 (-0.0009 to -0.0006) | <0.001 | <0.001 |
| FGF-2 | 0.0039 (0.0032 to 0.0045) | 0.0031 (0.0025 to 0.0037) | -0.0008 (-0.0016 to 0.0001) | 0.097 | 0.097 |
| GDNF | 0.0167 (0.0145 to 0.0189) | 0.0121 (0.01 to 0.0141) | -0.0047 (-0.0076 to -0.0018) | 0.002 | 0.004 |
| NGF-β | 0.0043 (0.0036 to 0.0050) | 0.0035 (0.0028 to 0.0042) | -0.0008 (-0.0017 to 0.0001) | 0.094 | 0.097 |
| Sensitive analysis: Normalized to 1 ml plasma |  |  |  |  |  |
| BDNF | 9.84 (8.16 to 11.52) | 8.65 (7.1 to 10.2) | -1.19 (-3.41 to 1.03) | 0.298 | 0.298 |
| EGF | 3.98 (3.62 to 4.34) | 2.01 (1.67 to 2.34) | -1.97 (-2.45 to -1.5) | <0.001 | <0.001 |
| FGF-2 | 13.94 (10.56 to 17.32) | 21.39 (18.27 to 24.51) | 7.45 (2.98 to 11.92) | 0.002 | 0.002 |
| GDNF | 60.55 (53.47 to 67.64) | 82.77 (76.24 to 89.31) | 22.22 (12.85 to 31.59) | <0.001 | <0.001 |
| NGF-β | 15.42 (11.71 to 19.13) | 24.56 (21.14 to 27.98) | 9.14 (4.23 to 14.05) | <0.001 | 0.001 |

Following ECT, there was a significant increase in EGF levels, this effect was also consistent across different normalization methods (**Table 2** and **Figure 2**). In summary, among the five neurotrophic factors tested, EGF showed the most robust results both at baseline and in terms of changes before and after ECT.

**Table 2.** Changes of neurotrophic factors in ADEVs of TRD after ECTs.

| Variable | Change from baseline (95% CI)) | Cohen's *d* (95% CI)) | *p* | *p<sub>FDR</sub>* |
| --- | --- | --- | --- | --- |
| Main analysis: Normalized to CD81 ratio |  |  |  |  |
| BDNF | 0.00001 (-0.00022 to 0.00024) | 0.01 (-0.33 to 0.36) | 0.934 | 0.934 |
| EGF | 0.00027 (0.00019 to 0.00036) | 1.14 (0.69 to 1.57) | <0.001 | <0.001 |
| FGF-2 | -0.00047 (-0.00118 to 0.00024) | -0.23 (-0.58 to 0.11) | 0.189 | 0.321 |
| GDNF | 0.00066 (-0.00101 to 0.00233) | 0.14 (-0.2 to 0.48) | 0.424 | 0.53 |
| NGF-β | -0.00048 (-0.00122 to 0.00026) | -0.23 (-0.58 to 0.12) | 0.193 | 0.321 |
| Sensitive analysis: Normalized to 1 ml plasma |  |  |  |  |
| BDNF | -1.31 (-3.03 to 0.41) | -0.27 (-0.61 to 0.08) | 0.131 | 0.131 |
| EGF | 1.2 (0.74 to 1.67) | 0.91 (0.5 to 1.31) | <0.001 | <0.001 |
| FGF-2 | -5.7 (-9.63 to -1.76) | -0.51 (-0.87 to -0.15) | 0.006 | 0.007 |
| GDNF | -9.15 (-15.48 to -2.82) | -0.51 (-0.87 to -0.15) | 0.006 | 0.007 |
| NGF-β | -6.74 (-11.19 to -2.29) | -0.54 (-0.9 to -0.17) | 0.004 | 0.007 |

### Causal influence of brain-specific *EGF* gene expression on depression

Given the significant correlation between EGF expression and depression above, we employed MR to investigate the causal influence of brain-tissue specific *EGF* and *EGFR* genes expression on depression. Adhering to MR assumptions, we identified cis-eQTL data for *EGF* and *EGFR* genes in the brain cortex, respectively (**sTable 2**). Our MR analysis suggests that decreased expression of the *EGF* gene in the cortex may be a potential risk factor for depression (Wald ratio OR= 0.95, *p* = 0.009) (**sFigure 3**). However, neither method found a significant causal relationship between *EGFR* and depression.

**Figure 3.**
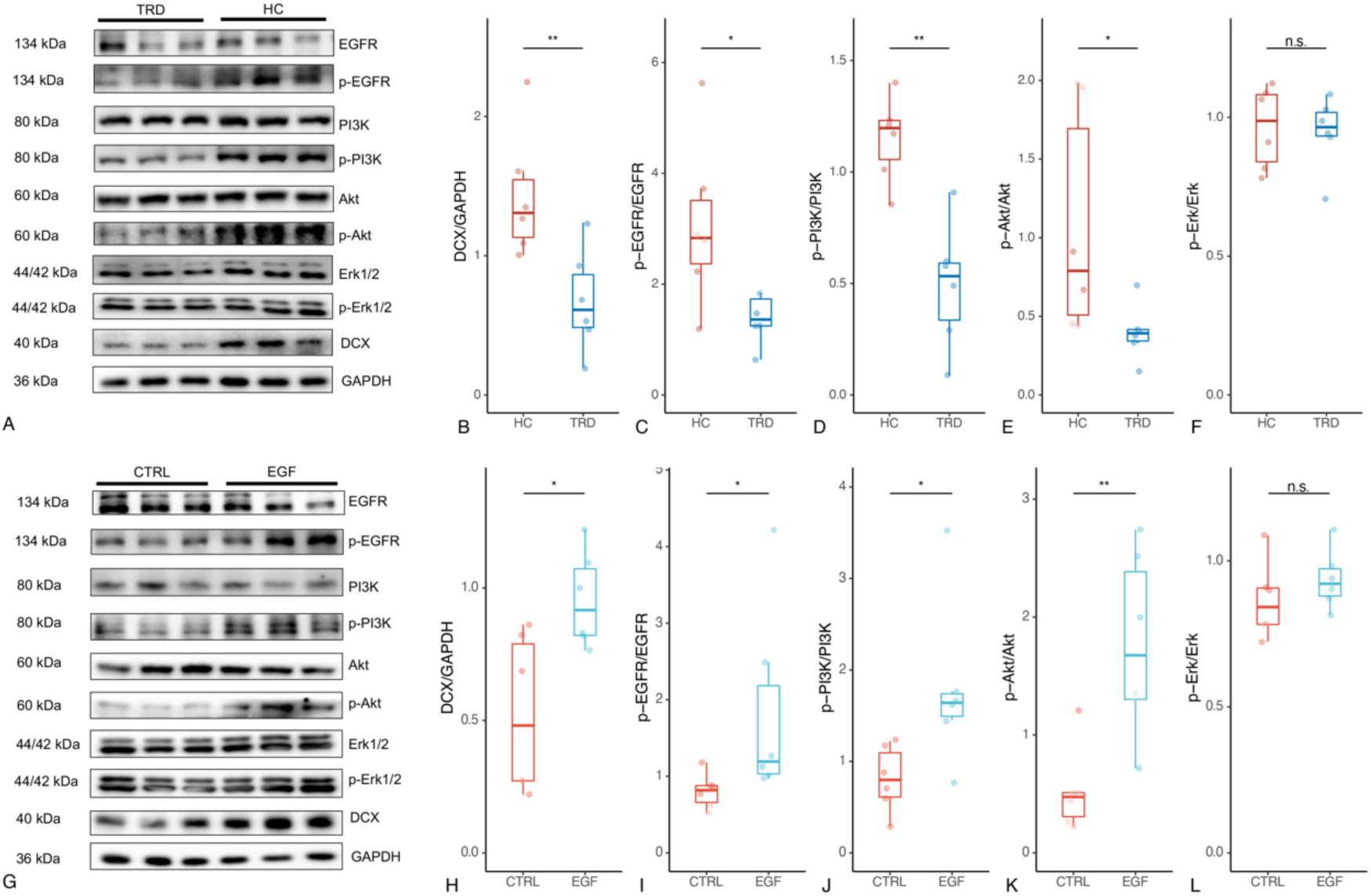
Western blotting results of cell expriments. A-F: After 24 hours of intervention with ADEVs from patients with TRD and HCs in SH-SY5Y cells, the expression levels of doublecortin (DCX) protein in the HC group significantly increased compared to the TRD group. The phosphorylation levels of EGFR, PI3K, and Akt also significantly increased, while there was no significant difference in the phosphorylation levels of Erk between the two groups. G-L: After 24 hours of intervention with ADEVs from patients with TRD and HCs in SH-SY5Y cells, the expression levels of DCX protein in the HC group significantly increased compared to the TRD group. The phosphorylation levels of EGFR, PI3K, and Akt also significantly increased, while there was no significant difference in the phosphorylation levels of Erk between the two groups. Note: n.s.: not significant; \**p*_FDR_ < 0.05; \*\**p*_FDR_ < 0.01.

Subsequently, we attempted to identify potential mechanistic pathways between *EGF/EGFR* genes and depression through MELODI Presto^31^. The analysis revealed that potential intermediary factors between these genes and depression were primarily associated with metabolic processes, cellular function, and neurological disorders. Interestingly, the results suggest an overlap between the *EGF*-depression interaction and the serotonin transporter pathway, potentially implicating *EGF* as a target for antidepressant development (**sTable 3**).

### EGF activates EGFR to participate in the promotion of DCX expression

ADEVs purified using UC were confirmed via TEM, NTA, and western blotting validating successful ADEVs collection (**sFigure 4**).

As shown in **Figure 3A-F**, after 24 hours of intervention with ADEVs from patients with TRD and HCs, the expression level of the DCX protein in SH-SY5Y cells intervened by ADEVs in the HC group showed a significant increase compared to the TRD group (*p* = 0.009). Moreover, there was a significant increase in the phosphorylation level of the EGFR (*p* = 0.041), along with simultaneous increases in the phosphorylation levels of downstream pathway proteins PI3K (*p* = 0.004) and Akt (*p* = 0.015). However, there was no significant difference in the phosphorylation level of Erk between the two groups (*p* = 0.818).

Following a 24-hour intervention with recombinant human EGF (**Figure 3G-L**), the expression level of the DCX protein in SH-SY5Y cells significantly increased compared to the control group (*p* = 0.041), accompanied by a significant elevation in the phosphorylation level of EGFR (*p* = 0.015). Furthermore, the phosphorylation levels of downstream pathway proteins PI3K (*p* = 0.015) and Akt (*p* = 0.004) were also significantly increased concurrently. Nonetheless, there was no significant difference in the phosphorylation level of Erk between the two groups (*p* = 0.240).

## Discussion

Our study, using plasma ADEVs, indicates that decreased levels of EGF in the CNS could serve as a biomarker for depression. Subsequently, MR results suggest that decreased expression of the *EGF* gene in the cortex may contribute to the development of depression. We also discovered a positive correlation between EGF levels in ADEVs and DCX, suggesting that the EGF/EGFR signaling pathway may play a role in regulating early neurogenesis traits in depression.

This study’s strength lies in directly acquiring CNS evidences through ADEVs and MR based on brain-tissue data, bypassing limitations of peripheral sources. Neurotrophic factors, like EGF, are often found at higher levels in peripheral tissues compared to the brain^17,18^. This widespread expression can confound results when solely using peripheral measures, a common limitation in depression studies. Additionally, the blood-brain barrier further complicates assessing CNS neurotrophic factors^32^. Therefore, our study offered valuable insights into the brain-specific relationship between neurotrophic factors and depression.

### EGF and depression

Previous research on EGF and depression has yielded conflicting results. While one study found elevated EGF levels in adolescents with depression or bipolar disorder^33^, another study focusing on elderly patients did not detect differences compared to HCs^34^, consistent with another case-control study^35^. Additionally, two studies reported decreased peripheral blood EGF levels in depressed patients^13,36^, consistent with findings of downregulated *EGF* mRNA levels in peripheral blood^37^. Additionally, cognitive alterations in MDD patients have been associated with the SNP rs2250724 of *EGF* gene^38^.

Heterogeneity in EGF findings likely stems from several factors. First, variations in study populations can influence results. Serum EGF levels demonstrably decline with age^39^, highlighting potential age-related heterogeneity. Second, comorbid conditions like anxiety might confound EGF expression. A study reported elevated EGF in patients with mild to moderate depression and anxiety^40^, while others found decreases in depression without anxiety^13,36^. Most importantly, previous research has primarily focused on assessing EGF levels in peripheral blood, where they can be influenced by various tissues^17^. This highlights the importance of directly assessing EGF expression within the CNS, precisely the approach employed in our study. We utilized ADEVs for the first time to detect EGF within the brains of depression patients, and observed a significant decrease in EGF levels in TRD patients compared to HCs. ROC curve analyses revealed high sensitivity and specificity of EGF in ADEVs in identifying TRD, suggesting it as a promising biomarker for depression.

Our study further revealed that levels of EGF in ADEVs increased following ECTs in patients with depression. Earlier studies have observed acute changes in EGF levels after ECT. For example, one study reported a significant decrease in serum EGF concentrations following ECTs^41^, while another focusing on schizophrenia showed no change in serum EGF levels between antipsychotic therapy alone and combined ECT and antipsychotic treatment^42^. Notably, these prior studies relied on peripheral EGF measurements, which may not reflect central EGF dynamics. However, our findings, based on ADEVs which directly reflect the CNS, suggest EGF as a potential marker for ECT response, highlighting the importance of CNS-level assessment for understanding and treating depression.

Additionally, our MR analysis indicated that genetically predicted decreases in *EGF* gene expression constitute a potential risk factor for depression, further supporting our clinical observations. As eQTL effects on gene expression can vary depending on the tissue examined, potentially introducing confounding factors and leading to incorrect inferences. To address this limitation, we specifically investigated the causal relationship between *EGF* eQTLs derived from brain-tissue specifically in the cortex and depression. To strengthen our results, we used the MELODI Presto analysis tool to further prioritize the *EGF* gene, highlighting its potential interactions with antidepressive agents. This provides additional evidence supporting the involvement of *EGF* in the mechanisms of antidepressant activity. This approach offers insights into the biological pathways linking *EGF* and depression, indicating that changes in the *EGF* expression could play a crucial role in the pathogenesis of depression.

The observed genetic associations with reduced *EGF* expression reinforce the need for additional preclinical studies to investigate how *EGF*-based interventions might benefit patients with depression, and they also shed light on how *EGF*-related pathways contribute to the onset and treatment of depression.

### EGF/EGFR, neurogenesis and depression

Our previous study using NDEVs, firstly provided human *in vivo* evidence of reduced markers of early neurogenesis DCX in patients with TRD, and observed increased traits of immature neurons following ECT^23^. One potential mechanism linking neurogenesis impairments with depression is through the loss of neurotrophic factors and the disruption of their associated signaling pathways^5^. This study discovered a notable positive correlation between EGF levels in ADEVs and DCX levels in NDEVs. This finding suggests a potential link between EGF and early neurogenesis. Then, we validated that ADEVs from HCs, with higher EGF levels, increased DCX expression in SH-SY5Y cells compared to ADEVs from patients with TRD. Similarly, EGF protein alone also stimulated DCX expression in SH-SY5Y cells, potentially via the EGFR-PI3K-Akt pathway.

Astrocytes are the predominant cell type within the neurogenic niche, creating a specialized environment for neurogenesis^43^. Previous studies have found that astrocytes can secret EGF to increase neurogenesis^8,44^. EGF can promotes the differentiation, maturation, and survival of various neurons, exerting neuromodulatory effects on different types of neurons in the CNS^44^. The exogenous EGF has specific effects on precursor cells *in vivo*^45^. Administering EGF into the ventricles for 2 weeks expands precursor cell populations in the subventricular zone (SVZ), decreases the total number of newborn neurons reaching the olfactory bulb, and significantly increases the production of olfactory bulb astrocytes. Moreover, EGF increases the number of newborn cells in the striatum either by migrating SVZ cells or by stimulating local precursor cells, while also boosting the number of newborn glial cells and reducing newborn neurons^46^. These findings illuminate EGF’s role in neurogenesis, and emerging evidence suggests that neurogenesis may play a role in depression^47^.

However, there is limited research exploring the relationship between EGF and neurogenesis in depression. One study shown that FGF, EGF, BDNF, ketamine, melatonin and the ketamine/melatonin combination stimulate neurogenesis and cell cluster formation in cultured human olfactory neuronal precursors, thus offering insights into the mechanism underlying their antidepressant effects^48^. Once EGF binds to its receptor, EGFR, it forms a homodimer, which undergoes autophosphorylation in the intracellular kinase domain, activating signaling cascades within the cell and even in the nucleus^49^. Upon EGFR activation, the main downstream signaling pathways include the PI3K-Akt pathway, the Ras-Raf-MAPK-Erk pathway, and the JAK/STAT signaling pathway^29^. Studies have shown that the PI3K-Akt and MAPK-Erk signaling pathways play crucial roles in neurogenesis, with several therapeutic targets for depression acting on these pathways to promote neuronal growth and survival^27,28^. Our findings suggest that EGF might exert its effect on early neurogenesis in depression by specifically activating the EGFR, which in turn triggers the PI3K-Akt pathway. This activation pattern contrasts with the MAPK-Erk pathway, often implicated in other cell growth and differentiation processes, indicating that the selective activation of PI3K-Akt by EGF could be a distinctive mechanism through which early neurogenesis is influenced in the context of depression. ECT may stimulate this signaling pathway by promoting EGF secretion, leading to increased early neurogenesis and potentially achieving antidepressant effects.

### EV, a candidate key mediator in mediating EGF-neurogesis signaling

During the process of EGF/EGFR signaling, EVs may play a significant role. EVs derived from neurons and neuroglial cells exert crucial bidirectional regulatory effects in the physiological and pathological processes of the CNS^50^. These nanoscale messengers not only ferry information from neurons to glial cells but also mediate the reverse pathway, influencing neuronal function and establishing a dynamic intercellular network^20^. For instance, neurons can transfer miR-124b to astrocytes via EVs, enhancing the expression of glutamate transporter-1 in astrocytes, thereby regulating synaptic plasticity^51^. Additionally, ADEVs can enhance neuronal survival under ischemic and hypoxic conditions by delivering antiviral proteins^52^. Regarding our study, results of cell experiments suggest a possible antidepressant mechanism between astrocytes and neurogenesis, EGF carried by ADEVs activates the EGFR signaling pathway (including PI3K-Akt) in neurons, thereby promoting neuronal proliferation and growth.

This study proposes a novel framework for investigating CNS diseases in human subjects. By leveraging brain-derived extracellular vesicles (BDEVs), we can gain initial insights into the *in vivo* CNS state, facilitating the study of real-world pathological conditions. Furthermore, complementing BDEV analysis with *in vitro* cell experiments offers a comprehensive understanding of CNS biology. Additionally, the potential of EVs as drug delivery vehicles, with their ability to cross the blood-brain barrier and target specific cell types, opens exciting avenues for developing targeted therapeutic strategies for CNS diseases.

## Limitations

This study has several limitations. Firstly, the limited quantity of ADEVs restricted high-throughput omics analyses, also hindering the detection of additional neurotrophic factors. Additionally, plasma ADEVs represent the entire brain, precluding identification of specific neurotrophic factor source regions. Elucidating these regional origins is helpful for understanding their roles in depression. Secondly, for the MR study, the major GWAS data populations were predominantly European, necessitating further validation of these MR findings in other populations. Also, the limited number of instruments for cis-eQTL data from cortex tissue and other brain tissues to investigate their association with depression, warranting further examination in future studies. Furthermore, large-scale independent tissue-specific eQTL datasets and employing additional genetic methods like transcriptome-wide association studies^53^ to estimate gene expression effects on depression should be considered in future studies. We look forward to future research addressing this issue more specifically. Thirdly, for cell experiments, resource constraints necessitated using the SH-SY5Y cell line instead of primary neural stem cells, potentially compromising the accuracy of neurogenesis data. Future studies should prioritize neural stem cells for a more faithful representation of neurogenesis processes. Additionally, serum-containing media hindered precise control over EGF levels, potentially influencing our results. Employing serum-free media would enhance experimental control. Finally, the complex composition of ADEVs limited our ability to identify the specific pro-neurogenic molecule(s). Future studies isolating and analyzing individual ADEV components are necessary to pinpoint the molecules responsible for the observed increase in neurogenesis.

## Conclusions

This study provides *in vivo* evidence for a potential role of reduced CNS EGF levels in TRD patients, suggesting EGF as a promising biomarker. Then using MR, we revealed a causal influence of low brain-specific *EGF* gene expression on depression. Furthermore, our findings highlight the involvement of the EGF/EGFR signaling pathway in regulating early neurogenesis, a process implicated in depression. These insights may offer critical insights for developing novel antidepressant therapies targeting the EGF/EGFR pathway using EVs.

## Supporting information

sFigure 1

sFigure 2

sFigure 3

sFigure 4

sTable 1

sTable 2

sTable3

Supplementary materials.Detailed Methods

## Data Availability

All data produced in the present study are available upon reasonable request to the authors.

## Acknowledgements

This work was supported by grants from the National Natural Science Foundation of China (U21A20364). No funding was received from commercial organizations.

## Authors’ contributions

SXX conducted methodology, investigation, data curation, and contributed to the original draft and conceptualization. HL contributed to data curation and participated in manuscript review and editing. MMC and KL conducted investigation and data curation. LY and CW participated in manuscript review and editing. XHX provided conceptualization, performed formal analysis, visualization and supervision. ZL provided project administration, supervision, and acquired funding.

## Supplementary materials

**sFigure 1.**
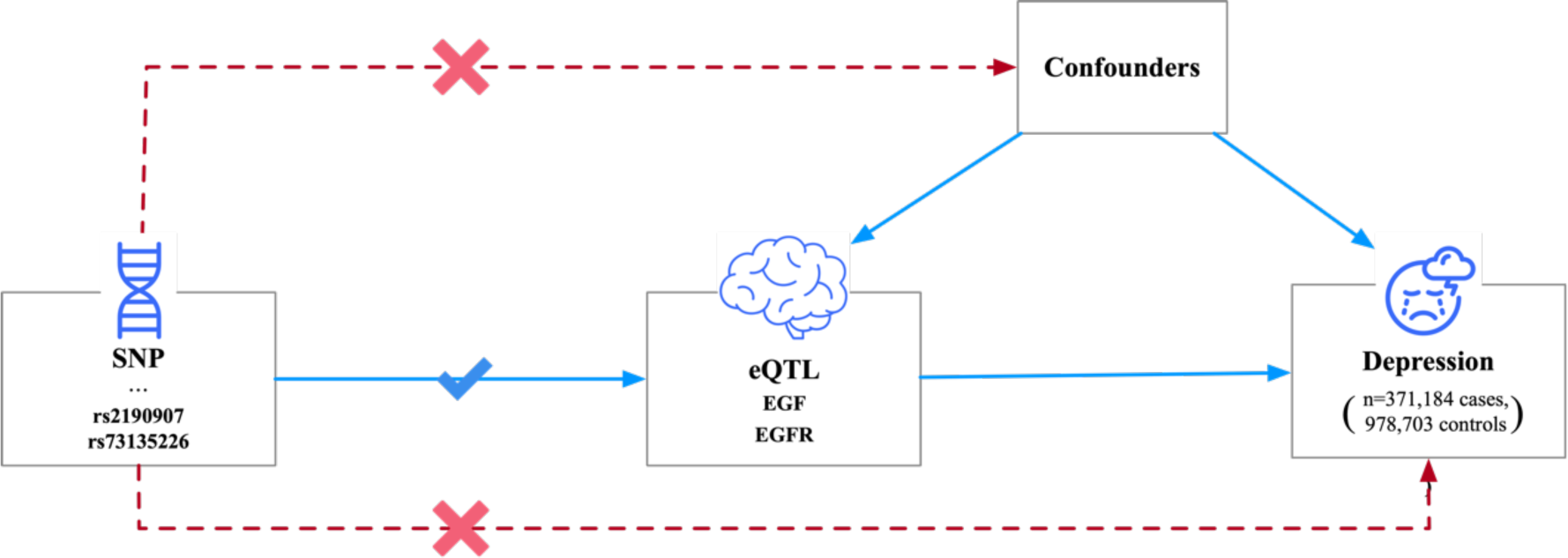
Mendelian randomization framework.

**sFigure 2.**
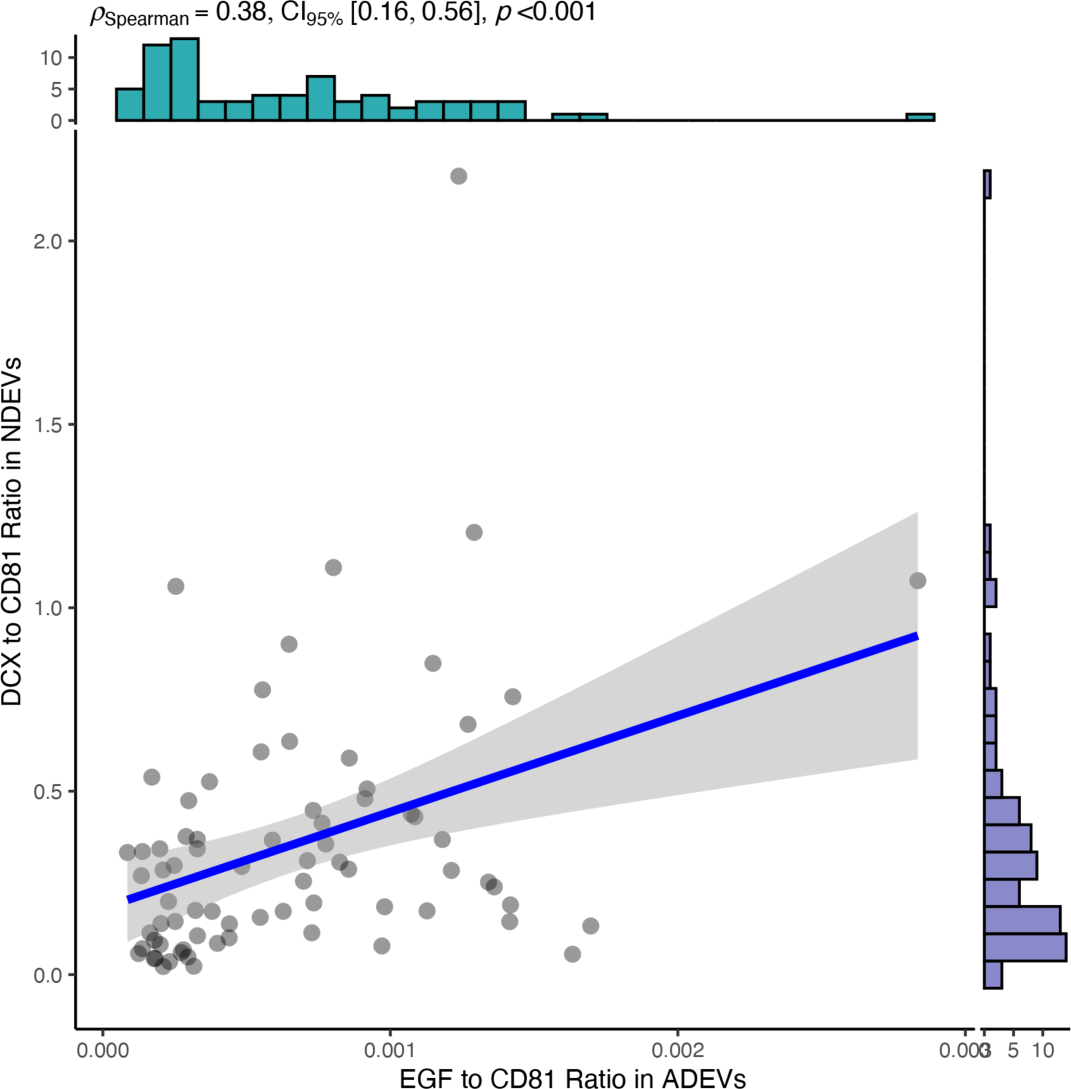
The spearman coefficient between EGF in ADEVs and the DCX in NDEVs.

**sFigure 3.**
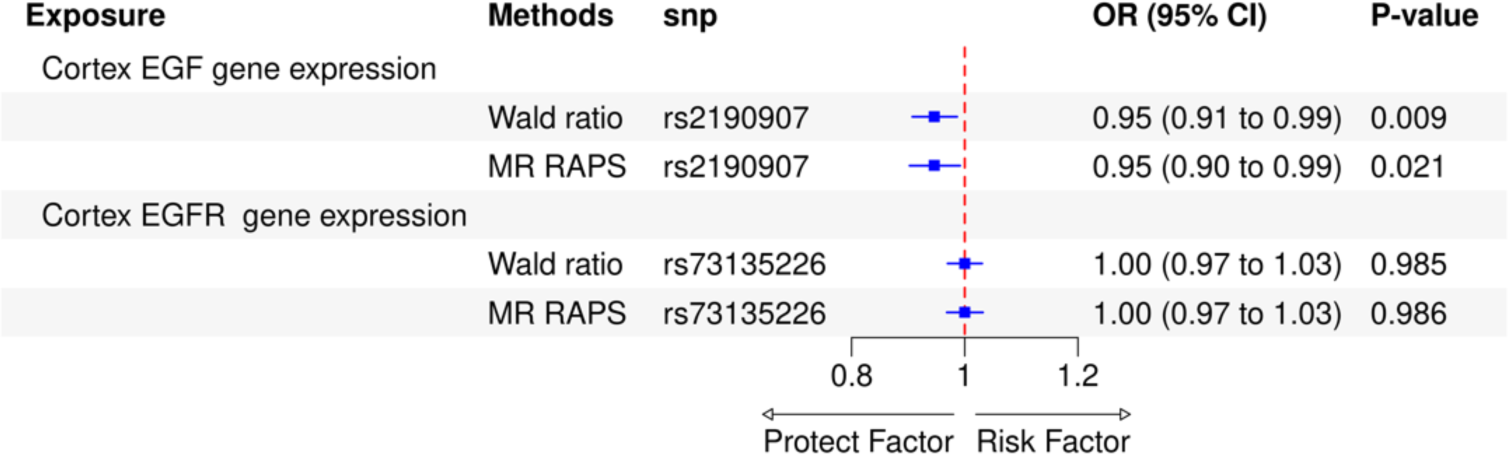
Results of Mendelian randomization study.

**sFigure 4.**
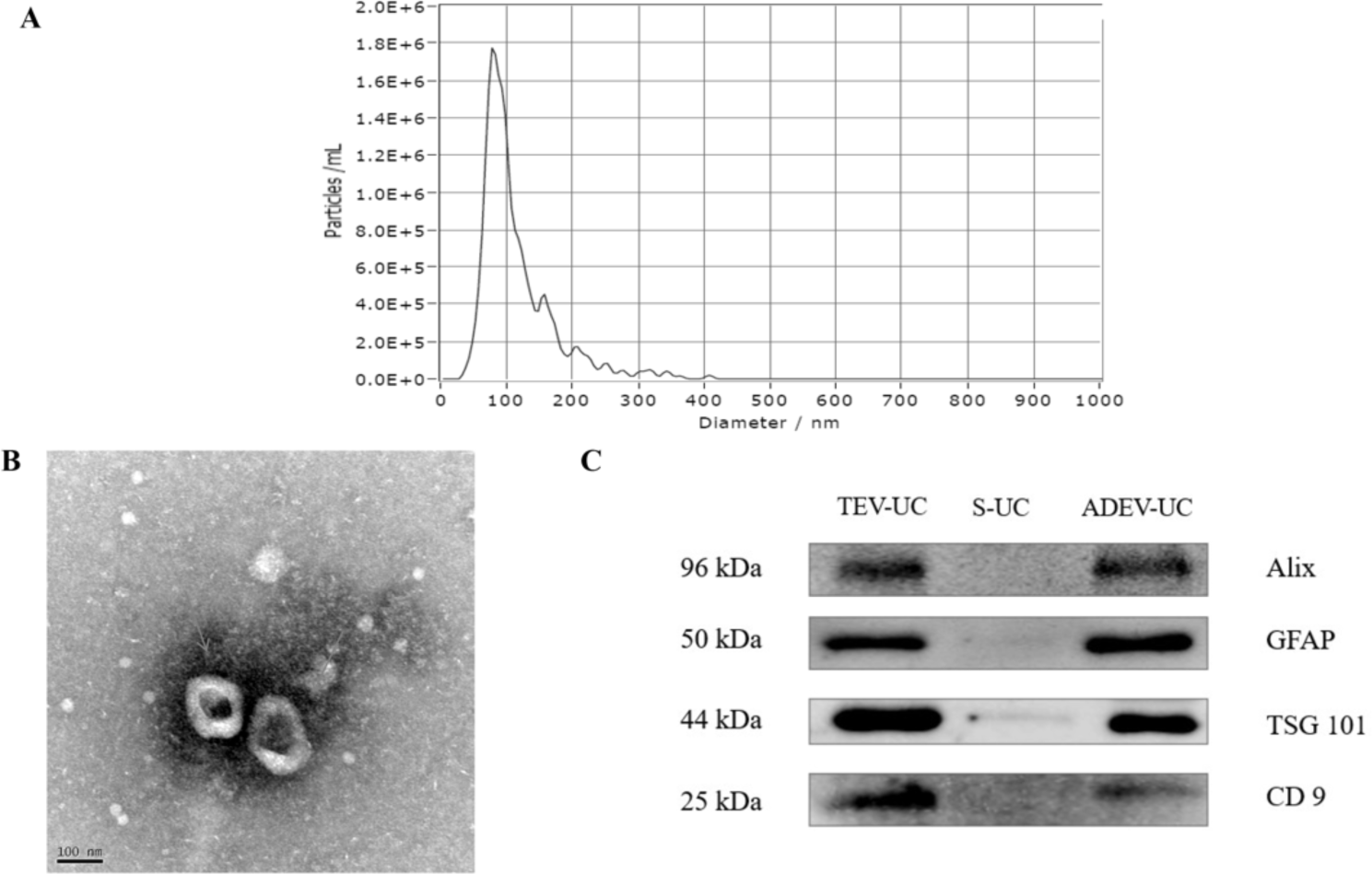
Confirmation of ADEVs utilizing UC. A) The results of NTA show the diameter distribution of the EVs, which also corroborate with the prior knowledge of exosomes-like small EVs (peak diameter: 82.3 nm, concentration: 2.4*10^9^ particles/ml); B) The transmission electron microscopy image of ADEVs isolated from a health control, the image clearly shows the exosomes-like shape (orange arrows); C) Western blotting results of total EV (TEV), supernatant of TEV, and ADEV from one healthy control. Three exosomes markers, CD9, Alix, TSG 101, and an astrocyte marker glial fibrillary acidic protein (GFAP) was identified.

**sTable 1.** Baseline characteristics of included participants for clinical trial.

**sTable 2.** Genetic instruments for *EGF/EGFR* expression in brain tissues.

**sTable 3.** MELODI Presto results for *EGF/EGFR* genes and depression.

**Supplementary document:** Detailed Methods.

