## Supplementary figures and images for "Epidermal Growth Factor in the Brain: A Promising Biomarker for Depression"

### sFigure 1

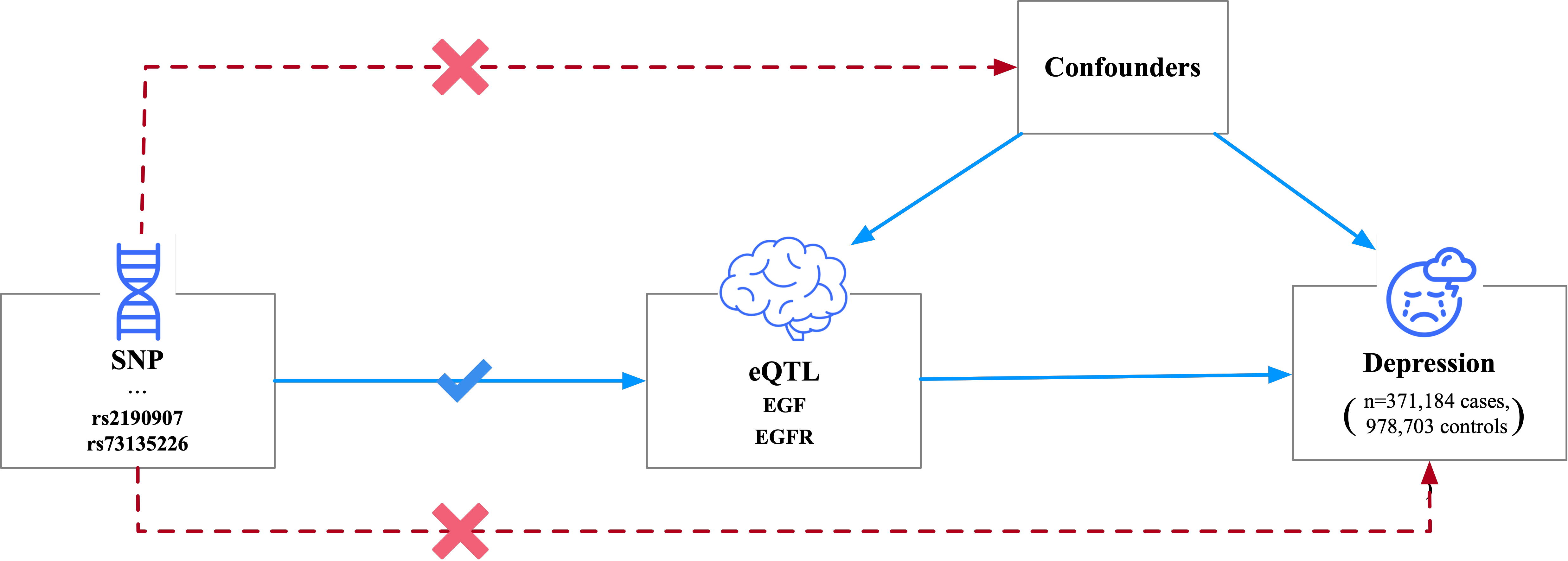

### sFigure 2

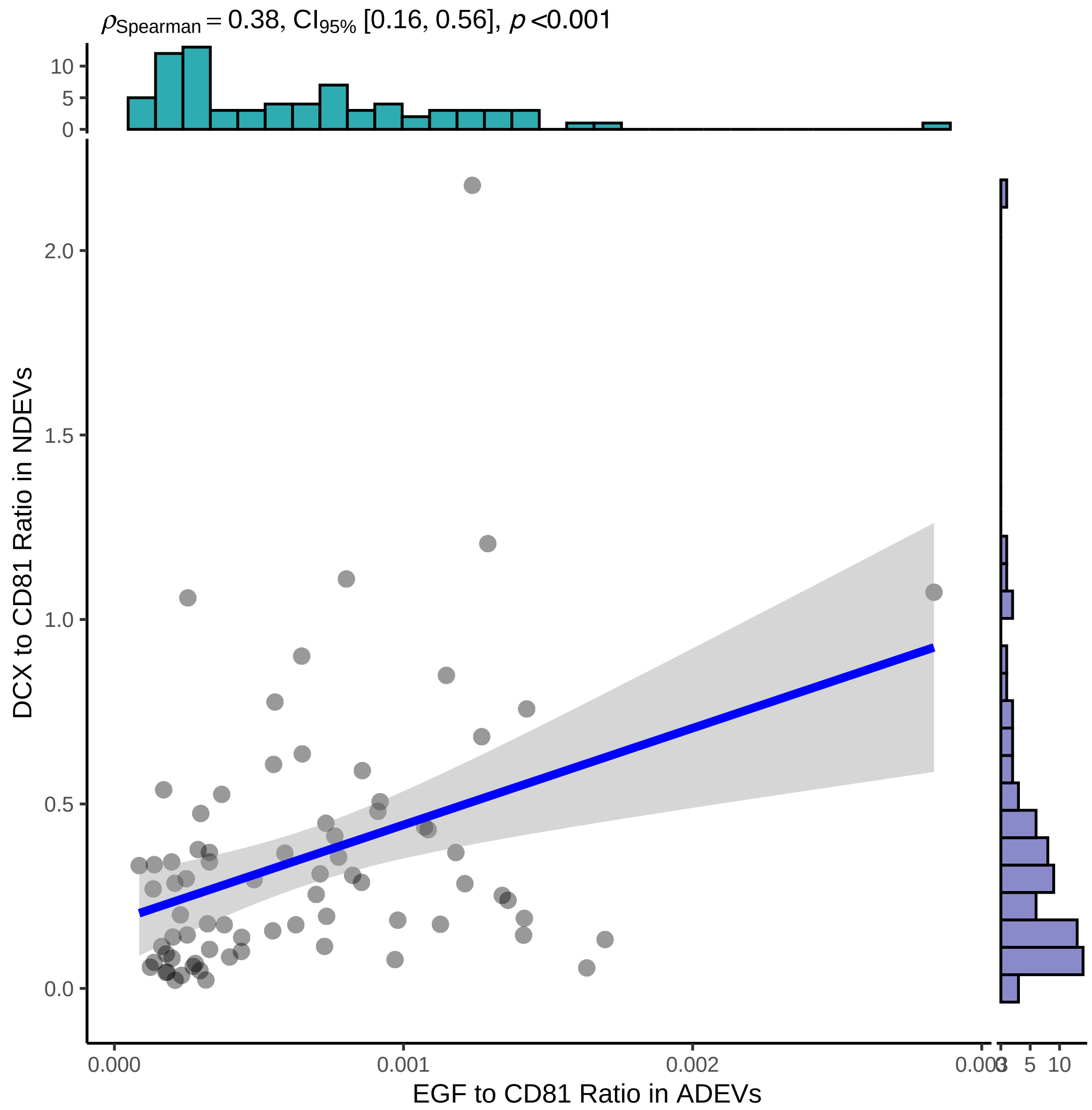

### sFigure 3

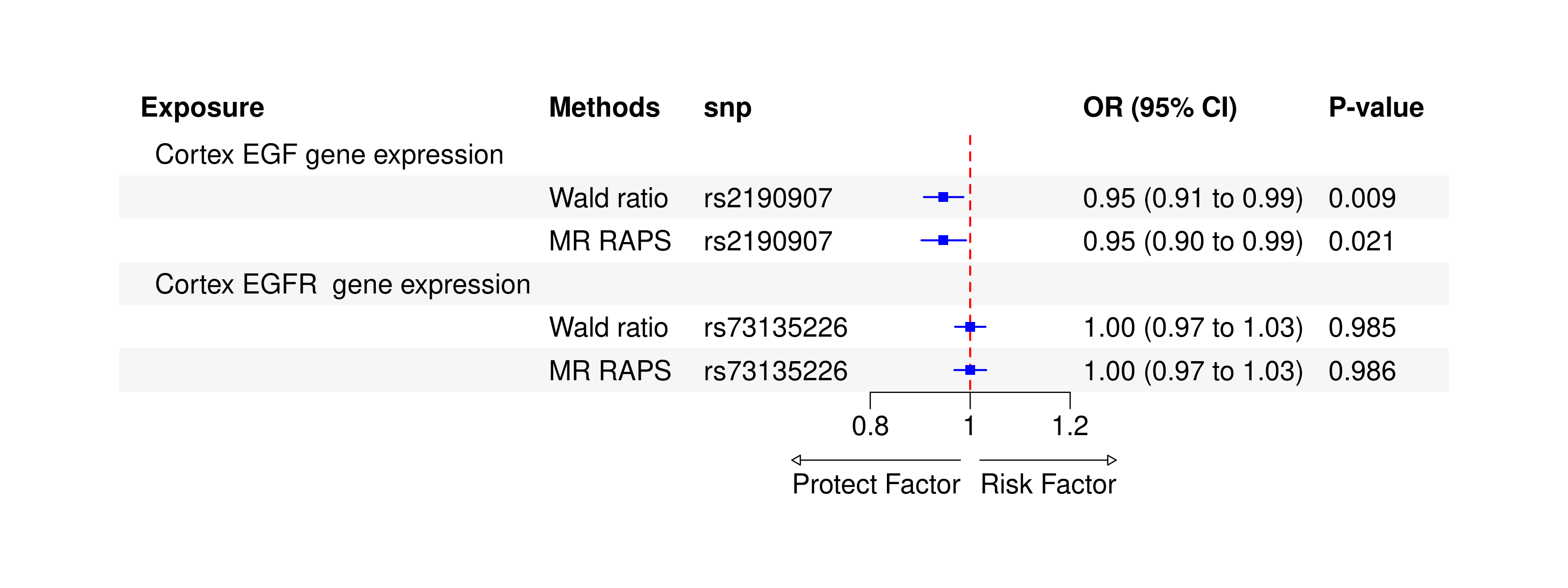

### sFigure 4

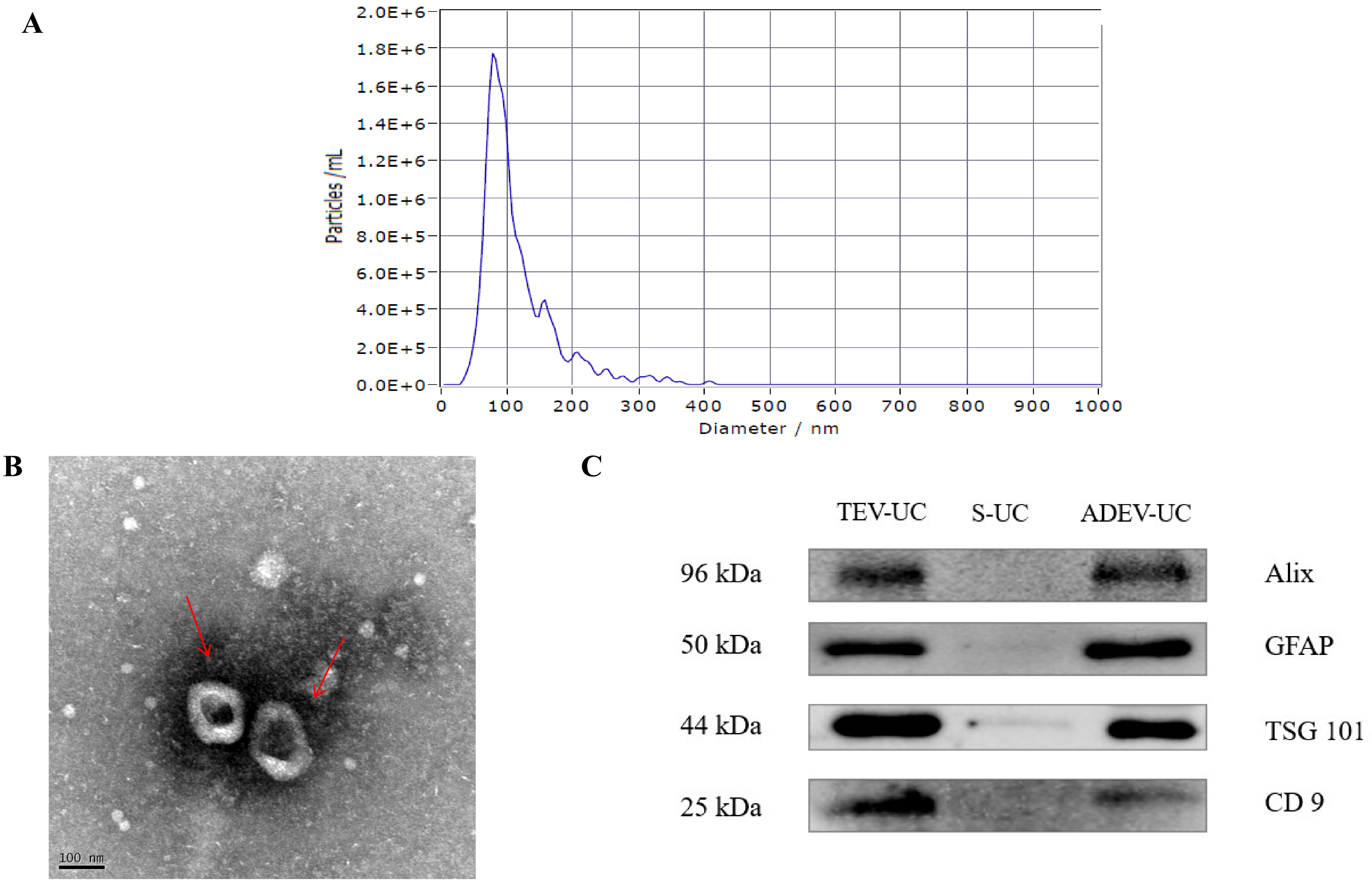
