## Supplementary material for "Epidermal Growth Factor in the Brain: A Promising Biomarker for Depression": sTable 1

sTable 1. Baseline characteristics of included participants for clinical trial.

| Variable | HC | TRD | *p* |
| --- | --- | --- | --- |
|  | (N = 35) | (N = 40) |  |
| Sex | |  | 0.640 |
| - Female | 23 (65.7%) | 24 (60.0%) | |
| - Male | 12 (34.3%) | 16 (40.0%) | |
| Age | 23.1 ± 3.9 | 21.8 ± 4.6 | 0.219 |
| EducationLevel | | | 0.272 |
| - PrimaryOrMiddle | 0 ( 0.0%) | 2 ( 5.0%) |  |
| - HighSchool | 23 (65.7%) | 22 (55.0%) | |
| - Undergraduate | 9 (25.7%) | 15 (37.5%) | |
| - Postgraduate | 3 ( 8.6%) | 1 ( 2.5%) |  |
| Education Years | 14.2 ± 1.3 | 13.6 ± 2.1 | 0.128 |
| Onset Age |  | 16.5 ± 4.6 |  |
| Disease Course/year |  | 5.4 ± 2.4 |  |
| History of ECT | | |  |
| - No |  | 37 (92.5%) | |
| - Yes |  | 3 ( 7.5%) |  |
| Ect Number |  | 6.1 ± 2.9 |  |
| Use of SSRIs | | |  |
| - No |  | 13 (32.5%) | |
| - Yes |  | 27 (67.5%) | |
| Use of SNRIs | | |  |
| - No |  | 28 (70.0%) | |
| - Yes |  | 12 (30.0%) | |
| Use of Other Antipresents | | |  |
| - No | 35 (100.0%) | 29 (72.5%) | |
| - Yes |  |  | |
| Fluoxetine Equivalent Dose/mg |  | 34.5 ± 21.4 |  |
| MADRS total score | 1.8 ± 2.2 | 36.8 ± 7.3 | <0.001 |
| MADRS: Cognitive Pessimism | 0.6 ± 1.0 | 17.2 ± 4.2 | <0.001 |
| MADRS: Affective | 0.5 ± 0.7 | 11.8 ± 2.4 | <0.001 |
| MADRS: Cognitive Anxiety | 0.2 ± 0.6 | 6.3 ± 2.5 | <0.001 |
| MADRS: Vegetative | 0.6 ± 0.9 | 5.9 ± 2.4 | <0.001 |
| RBANS total score | 506.1 ± 38.9 | 458.5 ± 47.3 | <0.001 |
| RBANS: Immediate Memory | 92.2 ± 13.2 | 81.3 ± 15.4 | 0.003 |
| RBANS: Visuospatial/Constructional | 104.8 ± 10.3 | 102.5 ± 12.3 | 0.395 |
| RBANS: Language | 101.2 ± 10.3 | 92.7 ± 11.6 | 0.002 |
| RBANS: Attention | 111.9 ± 13.2 | 97.8 ± 14.7 | <0.001 |
| RBANS: Delayed Memory | 96.0 ± 11.2 | 84.3 ± 16.1 | 0.001 |
