## Supplementary materials.Detailed Methods for "Epidermal Growth Factor in the Brain: A Promising Biomarker for Depression"

**ECT procedures**

Participants underwent a comprehensive pre-electroconvulsive therapy (ECT) assessment, including physical examination, laboratory tests, electrocardiogram, electroencephalogram, and evaluation for anesthesia and ECT risks. All individuals showed no clinically significant abnormalities. ECT was administered three times per week with bitemporal electrode placement using a Thymatron IV device (Somatics, LLC). Seizure threshold was determined at the first ECT session, starting at a dose level of 50 mC (or 10% of machine energy), and titrated upwards till a seizure of at least 15 seconds was induced. Subsequent ECT treatments were administered at 1.5 times seizure threshold. General anesthesia was induced using propofol (approximately 2 mg/kg), and myorelaxation with succinylcholine (approximately 1 mg/kg) and atropine (0.5 mg) before each session. Dosages were adjusted as needed. Discontinuation of ECT was decided by the psychiatrist based on factors such as reduced benefit, severe side effects, completion of 12 sessions, patient preference, and medical considerations. Patients could continue their prescribed antidepressants and antipsychotics, while anticonvulsant drugs, mood stabilizers, and benzodiazepines were discontinued throughout the ECT course.

**Blood samples**

Briefly, 6 ml blood was collected into BD Vacutainer tubes (BD Biosciences, Catalog# 367863) containing 10.8mg of K2 EDTA by using 0.7 mm needle from each participant. After collection, tubes were inverted 10 times immediately for proper mixing with anticoagulant. Tubes were transported vertically at room temperature without agitation. And then, the blood samples were centrifuged at 2,000 g for 15 minutes at room temperature to extract plasma within 2 hours of blood collection. Plasma was stored at −80°C until further use, and freeze-thawing was avoided as much as possible after that.

**Isolation of total extracellular vesicles (EVs)**

Total EVs were isolated from thawed plasma by the conventional differential ultracentrifugation (UC) and commercially available isolation kit ExoQuick (System Biosciences, CA, USA).

**Differential UC**

One-thousand μl of plasma was centrifuged at 10,000 g for 30 minutes at 4°C to remove contaminating debris. Then the supernatant diluted to a 4-ml Ultra-Clear tube (Beckman Coulter, Catalog# 344062) with phosphate-buffered saline (PBS, Beyotime Biotechnology, Shanghai, CN) was centrifuged at 100,000 g for 180 minutes at 4°C in a Beckman Optima XE-100 Ultracentrifuge using an SW 60 Ti rotor. Afterwards, the supernatant was discarded and total EVs were resuspended in 350 μl calcium and magnesium-free Dulbecco’s phosphate-buffered saline (DPBS, Beyotime) with protease and phosphatase inhibitor cocktails (PPIC, Beyotime) and was put at a rotating mixer for 2 hours at 4 ℃.

**ExoQuick isolation kit (ExoQuick)**

Briefly, 2 μl of thrombin (Beyotime, 611 U/ml) was added into 250 μl plasma to a final concentration of 5 U/ml. Then the sample was incubated at room temperature for 5 minutes while gently flicking the tube. After thrombin treatment, the sample was centrifuged at 10,000 g for 5 minutes at room temperature and the supernatant was transferred to a clean tube. It was stated by the manufacturer that “When isolating exosomes from plasma, fibrinogen and fibrin can impede efficient recovery. By pre-treating plasma with thrombin, the fibrinogen can be converted to fibrin and easily pre-cleared by a simple centrifugation step”. Then the plasma was gently mixed with 250 μl DPBS with PPIC at 4 ℃. Next, 0.5 ml of the obtained plasma was centrifuged at 3,000 g for 30 minutes. The supernatants were then mixed with 126 μl ExoQuick (Catalog# EXOQ20A-1) and incubated for 60 minutes at 4 ℃. After centrifugation at 1,500 g for 30 minutes, the total EVs precipitation was resuspended in 350 μl DPBS with PPIC and was put at a rotating mixer for 2 hours at 4 ℃.

**Isolation of astrocyte-derived extracellular vesicles (ADEVs)**

Each total EV sample was mixed with 50 μl 3% bovine serum albumin (BSA, Beyotime) and then incubated for 1 hour at room temperature with 2 μl Anti-GLAST (ACSA-1)-Biotin antibody (Miltenyi Biotec, Auburn, CA). Then, 10 μl of streptavidinagarose resin (Thermo Fisher Scientific, MA, US) and 40 μl of 3% BSA was added, and incubated for 30 minutes at room temperature. Then, after centrifugation at 800 g for 10 minutes at 4 ℃ and removal of the supernatant, each sample was resuspended in 100 μl cold 0.05M glycine-HCl (pH = 3.0) by gently mixed for 10 seconds and was centrifugated at 4,000 g for 10 minutes and recover the supernatant. Then mixed with 25 μl DPBS containing 3% BSA, and 10 μl 1M Tris-HCl (pH = 8.0, Beyotime) was added to adjust the pH to 7.0. The samples were then incubated for 10 minutes at 37 ℃ and was obtained as an ADEV sample. An aliquot of 20 μl was removed from each tube for ADEV counts before addition of 310 μl of mammalian protein extraction reagent (M-PER, Thermo Fisher Scientific) with PPIC.

**Transmission electron microscopy (TEM)**

Twenty μl of the ADEV sample was added dropwise to 200-mesh grids and incubated at room temperature for 10 minutes, then the grids were negatively stained with 2% phosphotungstic acid for 3 min, and the remaining liquid was removed by filter paper. Then observed with a HT7800 transmission electron microscope (Hitachi High-Tech Corporation, Tokyo, Japan).

**Nanoparticle tracking analysis (NTA)**

We measured the particle size and concentration using NTA with ZetaView PMX 110 (Particle Metrix, Meerbusch, Germany) and corresponding software ZetaView 8.04.02. Isolated ADEV samples were appropriately diluted using 1X PBS buffer (Biological Industries, Israel) to measure the particle size and concentration. The following parameters were set: Sensitivity: 80; Shutter: 100; Minimum area: 10 pixels; Maximum area: 1000 pixels; Minimum brightness: 30.

**Western blotting for ADEVs confirmation**

The protein concentration of EVs preparations was measured using a BCA protein assay kit (Beyotime) in accordance with the manufacturer’s instructions. The total protein content was calculated by multiplying the protein concentration by the volume of the isolated EVs.

For each sample, protein was loaded onto a 12% polyacrylamide gel. Following electrophoresis, the proteins were transferred from the gel onto a polyvinylidene fluoride membrane (Millipore) using a Trans‐Blot Turbo Transfer System (Bio‐Rad). The membrane was blocked with 5% non‐fat dry milk in TBST for 30 minutes at room temperature and incubated with primary rabbit anti‐human of CD9 antibody (Abcam, Catalog# ab 263019), rabbit anti‐TSG101 antibody (Abcam, Catalog# ab125011), mouse anti-Alix antibody (Proteintech, Catalog #67715-1-Ig) and rabbit anti-glial fibrillary acidic protein (GFAP) antibody (Abcam, Catalog# ab68428) overnight at 4°C. After three washes with TBS-t buffer, membranes were incubated for 60 minutes at room temperature with HRP‐conjugated goat anti‐rabbit highly cross-adsorbed secondary antibody (Invitrogen, Catalog# A16110) and HRP-conjugated goat anti-mouse secondary antibody (Servicebio, Catalog# GB23301). Membranes were developed with Clarity enhanced chemiluminescence substrate (Servicebio, Wuhan, CN, Catalog# G2014-50ML), and the signal was acquired with Chemi-Doc MP (Bio-Rad Laboratories) and analyzed using Image Lab software (Bio-Rad Laboratories). Normalization was performed against total protein load signal.

**Mendelian Randomization (MR)**

**Study Design**

To investigate the potential causal association between *epidermal growth factor (EGF)*, *EGF receptor (EGFR)* and depression, we employed MR leveraging Genome-Wide Association Study (GWAS) summary data on depression and expression quantitative trait loci (eQTL) from brain tissues^[1]^. Within the MR framework^[1-3]^, eQTLs for *EGF* and *EGFR* derived from brain tissues, utilized as exposures, with corresponding Single Nucleotide Polymorphisms (SNPs) serving as instrumental variables (IVs), and depression as the outcome (sFigure 1). The MR approach rests on three critical assumptions^[4]^: 1) strong association between the SNPs and *EGF/EGFR* expression levels, 2) SNPs are independent of confounders influencing the *EGF*-depression association, and 3) the SNPs solely influence depression risk through their effect on *EGF/EGFR* expression.

**Data Sources and Instrument Variable Selection**

*EGF* and *EGFR* expression data were obtained from the MetaBrain consortium (<https://www.metabrain.nl/cis-eqtls.html>)^[5]^. This dataset harmonized RNA-sequencing data (8,613 samples) from 14 brain-related datasets and conducted cis- and trans- eQTL meta-analyses in multiple brain region-specific datasets, encompassing data from up to 2,683 individuals of European ancestry.

We selected SNPs at the stringent genome-wide significance threshold (*p* < 5 *10^-8^) as IVs. Specifically, one SNP (rs2190907) was identified as the IV for *EGF*, and four SNPs (rs73135226, rs6964933, rs74504435, rs7795458) served as IVs for *EGFR* (**sTable 2**).

GWAS summary data on depression was obtained from the study conducted by Thomas D. Als et al., as the largest GWAS meta-analysis of depression to date, including >1.3 million individuals (371,184 with depression) and identified 243 risk loci^[6]^. Due to data access restrictions, we excluded data from 23andMe, resulting in a final dataset of 294,322 cases and 741,438 controls.

**MR analyses**

MR analyses were performed using the *TwoSampleMR* R package (<https://mrcieu.github.io/TwoSampleMR/index.html>). Following the selection of eligible IVs, we implemented linkage disequilibrium (LD) clumping with an *R^2^*< 0.001 within a 10,000 kb window to minimize potential bias from pleiotropy. Next, the IVs were extracted from the outcome trait and were harmonized in both exposure and outcome GWAS datasets. MR effects were then estimated using the Wald ratio^[7]^ for analyses with a single independent IV and IVW^[2]^ with multiple IVs. Additionally, MR-Robust Adjusted Profile Score (MR-RAPS)^[8]^ was employed utilizes a profile-likelihood function to estimate the variance of pleiotropic effects, down-weighting outliers during causal estimation. Sensitivity analysis will be conducted with a sufficient number of IVs, comprising at least three IVs. We applied the Bonferroni correction to determine the adjusted significance threshold (*p* <0.05/2).

**MELODI Presto**

We conducted the literature review using text mining approaches and identified overlapping enriched elements between EGF/EGFR and depression. The literature information was obtained from SemMedDB, implemented in MELODI Presto^[9,10]^, which summarizes the literature information as “subject-predicate-object” triples. The object is the phenotype of interest, for instance, “Depression”. The predicate is a term linking the subject and object together, where the term “INTERACTS_WITH” implies interaction, “STIMULATES” implies stimulation, “CAUSES” implies causality, the term “ASSOCIATED_WITH” implies association, and the term “COEXISTS_WITH” implies co-existence. The enrichment odds ratio and *p* value for each triple was estimated based on the number of appearances in the literature (e.g. the number of PubMed IDs supporting the existence of the triple).

MELODI Presto detected 128 overlapping enriched elements between EGF/EGFR and depression items evidence from the scientific literature **(sTable 3)**. Among these, 89 interactions with Antidepressive Agents and 10 serotonin transporter overlap enrichment semantic triples, providing insights into the potential biological pathways and mechanisms connecting EGF/EGFR and depression.
